# Nephrotic Syndrome Disease Activity Predicts Severity of the Associated Hypercoagulopathy

**DOI:** 10.1101/2020.03.13.20035493

**Authors:** Amanda P. Waller, Jonathan P. Troost, Samir V. Parikh, Katelyn J. Wolfgang, Brad H. Rovin, Marvin T. Nieman, William E. Smoyer, Matthias Kretzler, Bryce A. Kerlin, for The NEPTUNE Investigators

## Abstract

Nephrotic syndrome (NS) is associated with an acquired hypercoagulopathy and strong predilection for life-threatening thrombotic complications. Current anticoagulant prophylaxis guidelines are based upon controversial hypoalbuminemia thresholds. Anticoagulant prophylaxis is thus inconsistently implemented due to a lack of high-grade safety and efficacy data. Development of evidence-based clinical parameters that define thrombosis risk may thus refine safe and effective anticoagulant use. Endogenous thrombin potential (ETP) is a recognized measure of hypercoagulopathy and established predictor of both incident and recurrent thrombosis. This study utilized biorepository samples from a prospective longitudinal cohort study to demonstrate that ETP is proportional to NS disease activity, resulting in multivariable models that are significantly correlated with ETP. The relationship with disease activity was confirmed in a separate cohort. These models revealed that ETP is related to disease activity in a manner dependent on remission status and that proteinuria and hypercholesterolemia exert the strongest influence on ETP. In contrast to prior epidemiology studies, which did not include hypercoagulopathy measures, we found that hypoalbuminemia was less predictive of ETP. These findings are consistent with our previously reported animal model observations and are expected to inform the design of clinical trials that will generate high-grade evidence to guide more effective and safer anticoagulant use and thus reduce life-threatening thrombotic events in patients with NS.

## Introduction

Nephrotic syndrome (NS), a leading cause of end stage kidney disease (ESKD), is characterized by massive proteinuria, hypoalbuminemia, edema, and hyperlipidemia.^1-3^ As recently reviewed by our group and others, venous thromboembolism (VTE) is a frequent life-threatening complication of NS.^4-7^ Up to 27% of adults and 3% of children with NS develop VTE during their disease course, with the majority of VTE occurring within 90 days of diagnosis.^5, 8^ The frequency of VTE in adult NS is thus higher than both the well-known cancer-VTE predilection (≤5.7%) or the medical hospitalization VTE-rate (^∼^20%), but less than the ^∼^40% VTE-incidence amongst surgical admissions.^9^ Therefore, although NS is a rare disease, accounting for only a small proportion of total VTE cases, it nonetheless carries a profound VTE risk. Arterial thrombosis is less common than VTE in adult NS and is rarely reported in childhood NS.^10-12^ NS-associated VTE risk is secondary, at least in part, to the well-known acquired hypercoagulopathy that develops during NS (NS-hypercoagulopathy).^4-6^

While VTE complications may be preventable with prophylactic anticoagulation, this intervention remains controversial in NS due to a lack of well-defined indications and randomized controlled trial data demonstrating safety and efficacy.^8, 13-16^ For example, current KDIGO guidelines cite low-grade evidence, based upon Markov modeling of a virtual NS cohort, to suggest prophylactic anticoagulation when serum albumin is <2.5 g/dL.^15^ Recent epidemiologic studies have revealed that both proteinuria and hypoalbuminemia are associated with the likelihood of VTE, with higher proteinuria and lower serum albumin correlating with VTE risk, strongly suggesting that NS disease activity may be proportional to the severity of hypercoagulopathy.^10, 11, 17-20^ Thus, rigorous determination of the relationship between hypercoagulopathy and NS activity biomarkers offers the opportunity to identify clinically meaningful thresholds which may be implemented in clinical trials designed to generate high-grade evidence regarding the safe and effective use of prophylactic anticoagulation for NS patients.

Recently, thromboelastography, which determines the viscoelastic properties of clotting whole blood, was demonstrated to increase proportionally to NS activity.^21^ Similarly, *ex vivo* plasma thrombin generation has been used to show that thrombin generation is elevated in NS patients.^22^ Meanwhile, our group recently reported that both proteinuria and hypoalbuminemia are proportional to hypercoagulopathy, as assessed by *in vivo* thrombosis, viscoelasticity, and thrombin generation in rat NS models.^23^ Both thromboelastography and thrombin generation are ‘global hemostasis’ assays.^24^ The latter measures thrombin production over time, including the period after fibrin clot formation (which is the end point of conventional clinical coagulation assays) and is more amenable to use in centralized laboratories because frozen plasma can be used instead of whole blood.^24^ Thrombin is the key effector enzyme of the hemostatic system and, because of its negative and positive feedback loops, the amount of thrombin produced by the hemostatic system is a product of both upstream and downstream coagulation cascade status.^25, 26^ Several parameters are derived from the assay, including endogenous thrombin potential (ETP), which represents the area under the thrombin activity-time curve. Importantly and critical for this study, ETP is a biomarker of hypercoagulopathy that has demonstrated predictive value for both incident and recurrent VTE-risk in non-NS patient populations.^27-36^ The aim of the present study was thus to determine the relationship between ETP and NS activity biomarkers in human NS.

## Results

### Demographics and Clinical Characteristics

The <u>Nep</u>hrotic Syndrome S<u>tu</u>dy <u>Ne</u>twork (NEPTUNE) cohort consisted of plasma samples from 150 unique subjects randomly selected to represent the available proteinuria spectrum within the NEPTUNE biorepository as of October 2014. Three (2%) samples had undetectable thrombin generation indicating that the plasma had clotted during the pre-analytic phase; the remaining 147 samples were thus advanced for further analysis (**Table 1**). Patients with minimal change disease (MCD) or membranous nephropathy (MN) were significantly younger or older (median age 17 vs. 55 years, respectively) than those with focal segmental glomerulosclerosis (FSGS; median age 39 years; *p*<0.05). Patients with MCD (53%) or FSGS (32%) were more likely to be on immunosuppressive therapy (vs. 21% of MN patients; *p*<0.05). Urinary protein-to-creatinine ratio (UP:C) was significantly higher (median 3.5 g/g creatinine) in the MN patients (vs. 0.3 g/g and 1.9 g/g for MCD and FSGS, respectively; *p*<0.05). Because serum albumin (the conventional clinical laboratory method) is not uniformly determined on simultaneously collected serum specimens in a NEPTUNE central laboratory, we performed albumin measurements on the plasma aliquots as a surrogate method. Plasma albumin was similar in the MCD and FSGS groups (median 3.3 and 3.4 g/dl, respectively) and significantly higher than the MN group (2.5 g/dl; *p*<0.05). FSGS patients had the lowest estimated glomerular filtration rate (eGFR; 60 ml/min/1.73m^2^ vs. 81 and 97 for MN and MCD, respectively; *p*<0.05). The distributions of UP:C, plasma albumin, total cholesterol, and eGFR across all 147 patients are shown in **Figure 1**. Thrombin generation assays are susceptible to confounding by preanalytical variables, such as sample collection technique and processing, shipping, and freeze/thaw cycles, all of which are unknowns when using biorepository samples.^37^ Thus, to enable validation of ETP signals observed in the NEPTUNE cohort, we also collected urine and plasma from a second, locally-derived group of NS patients, for whom we were able to carefully control these preanalytical variables. This “Columbus cohort” consisted of 23 newly-diagnosed NS patients, the characteristics of which are shown in **Table S1**.

**Table 1.** Demographics and Clinical Characteristics of the NEPTUNE Cohort.

|  | All<br>(n=147) | MN<br>(n=28) | MCD<br>(n=45) | FSGS<br>(n=74) |
| --- | --- | --- | --- | --- |
| Age (years), median (IQR)* | 37 (15, 55) | 55 (42, 63) | 17 (11, 31) | 39 (16, 56) |
| Age (<18), n (%) | 45 (31) | 0 (0) | 23 (51) | 22 (30) |
| Female, n (%) | 60 (41) | 14 (50) | 23 (51) | 23 (31) |
| Race, n (%) |  |  |  |  |
| Multi-racial | 11 (7) | 1 (4) | 7 (16) | 3 (4) |
| Asian/Asian American | 14 (10) | 3 (11) | 5 (11) | 6 (8) |
| Black/African American | 41 (28) | 6 (21) | 10 (22) | 25 (34) |
| Native Hawaiian/Other Pacific Islander | 1 (1) | 0 (0) | 0 (0) | 1 (1) |
| White/Caucasian | 76 (52) | 17 (61) | 22 (49) | 37 (50) |
| Unknown | 4 (3) | 1 (4) | 1 (2) | 2 (3) |
| Hispanic Ethnicity, n (%) | 25 (17) | 5 (18) | 6 (13) | 14 (19) |
| Current Smoker, n (%) | 0 (0) | 0 (0) | 0 (0) | 0 (0) |
| Any alcohol use, n (%) | 59 (41) | 12 (43) | 13 (30) | 34 (47) |
| Treatment at time of collection, n (%) |  |  |  |  |
| Lipid modifying agents | 53 (36) | 19 (68) | 8 (18) | 26 (35) |
| Steroids | 44 (30) | 5 (18) | 19 (42) | 20 (27) |
| CNI | 18 (12) | 2 (7) | 8 (18) | 8 (11) |
| MMF | 3 (2) | 0 (0) | 2 (4) | 1 (1) |
| CTX | 3 (2) | 1 (4) | 1 (2) | 1 (1) |
| Rituximab | 2 (1) | 1 (4) | 1 (2) | 0 (0) |
| Any immunosuppression* | 54 (37) | 6 (21) | 24 (53) | 24 (32) |
| RAAS-Blockade | 71 (48) | 15 (54) | 15 (33) | 41 (55) |
| UP:C (g/g) , median (IQR)* | 1.9 (0.3, 3.6) | 3.5 (2.4, 5.2) | 0.3 (0.1, 1.9) | 1.9 (0.9, 3.2) |
| Plasma albumin (g/dl), median (IQR)* | 3.2 (2.4, 3.7) | 2.5 (2.1, 3.3) | 3.3 (2.6, 3.7) | 3.4 (2.5, 3.8) |
| eGFR (ml/min/1.73m<sup>2</sup>), median (IQR)* | 82 (50, 106) | 81 (51, 101) | 97 (83, 119) | 60 (44, 100) |
| Total Cholesterol (mg/dl), median (IQR)* | 230 (189, 280) | 255 (213, 328) | 207 (174, 265) | 235 (190, 278) |
| Other medical conditions (ever), n (%) |  |  |  |  |
| Heart Failure | 1 (1) | 0 (0) | 0 (0) | 1 (1) |
| Stroke | 4 (3) | 2 (7) | 0 (0) | 2 (3) |
| Cancer | 13 (9) | 5 (18) | 3 (7) | 5 (7) |
| Hypertension* | 91 (62) | 20 (71) | 19 (42) | 52 (70) |
| Diabetes | 16 (11) | 6 (21) | 2 (4) | 8 (11) |
| Heart arrhythmia | 10 (7) | 2 (7) | 4 (9) | 4 (5) |
| PVD | 2 (1) | 1 (4) | 0 (0) | 1 (1) |
| CAD | 6 (4) | 1 (4) | 0 (0) | 5 (7) |
| VTE | 4 (3) | 2 (7) | 1 (2) | 1 (1) |
| Rheumatological disease | 15 (10) | 4 (14) | 4 (9) | 7 (9) |
| Any thromboembolic event<sup>†</sup> | 13 (9) | 4 (14) | 1 (2) | 8 (11) |
| Before ETP sample collection<sup>‡</sup> | 10 (7) | 3 (11) | 1 (2) | 6 (8) |
| After ETP sample collection<sup>††</sup> | 4 (3) | 2 (7) | 0 (0) | 2 (3) |

MN: membranous nephropathy, MCD: minimal change disease, FSGS: focal segmental glomerulosclerosis. IQR: interquartile range. CNI: calcineurin inhibitor, MMF: mycophenolate mofetil, CTX: cyclophosphamide, RAAS: renin-angiotensin-aldosterone system. UP:C: urine protein-to-creatinine ratio, eGFR: estimated glomerular filtration rate. PVD: peripheral vascular disease, CAD: coronary artery disease, VTE: venous thromboembolism. <sup>†</sup>Stroke, PVD, CAD, or VTE. <sup>‡</sup>Patients with VTE prior to enrolment were excluded. <sup>††</sup>Evaluated through October 2019 (5 years). \*p<0.05 between MN vs. MCD vs. FSGS. Chi-square test for categorical; Kruskal-Wallis for continuous.

**Fig. 1.**
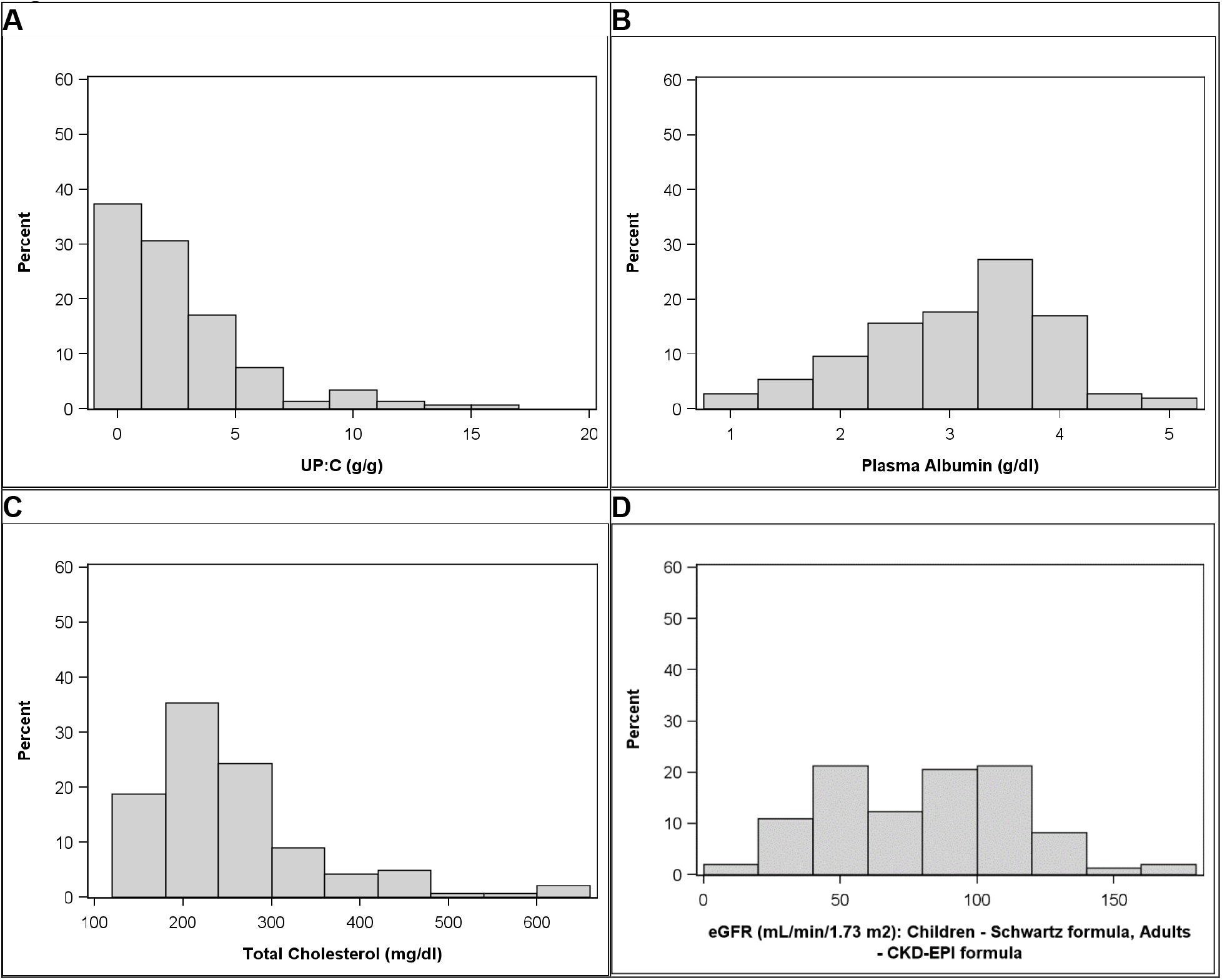
Clinical Characteristics of the NEPTUNE Cohort. Nephrotic syndrome disease activity distributions at the time of endogenous thrombin potential sample collection amongst the 147 NEPTUNE participants studied: (**A**) UP:C (urine protein-to-creatinine ratio), (**B**) plasma albumin, (**C**) total cholesterol, and (**D**) eGFR (estimated glomerular filtration rate).

### Univariate Relationships with Endogenous Thrombin Potential

To address the hypothesis that ETP is proportional to disease activity, we performed univariate analyses comparing ETP to markers of NS disease activity (e.g. UP:C, plasma albumin, lipids), various NS therapeutics, and known risk factors for cardiovascular disease (e.g. tobacco use, alcohol use, age) available in the NEPTUNE database. As shown in **Table 2**, ETP was significantly correlated with plasma albumin (Spearman’s rho (*ρ*) rank correlation coefficient *ρ*=‾0.31; *p*<0.001; **Figure 2A**) and urinary protein-to-creatinine ratio (UP:C; *ρ*=0.35; *p*<0.001; **Figure 3A**). ETP was also significantly correlated with total cholesterol (*ρ*=0.42; *p*<0.001; **Figure 2B**) and other components of the lipid profile (**Table 2**). ETP was also correlated with eGFR (*ρ*=0.17; *p*=0.04; **Figure 2C**). Age was negatively correlated with ETP (*ρ*=‾0.19; *p*=0.02; **Figure 2D**). However, this relationship was primarily driven by higher ETP in children 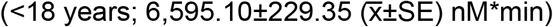 vs. adults (≥18 years; 5,705.90±160.79 nM*min; *p*=0.002) and ETP was not correlated with age within either age group sub-cohort. Treatment with steroids and renin-angiotensin-aldosterone system (RAAS) blockade were both correlated with ETP, but with opposing effects (**Table 2**). Whereas steroid therapy was positively correlated with ETP, RAAS-blockade was associated with decreased ETP. Alcohol use was negatively correlated with ETP, whereas tobacco use did not influence ETP in this study. Germane to our hypothesis, similar univariate relationships with UP:C and plasma albumin were identified in the Columbus cohort (**Table S2**). Unfortunately, there was insufficient power to assess hyperlipidemia and eGFR in the Columbus cohort because data for these unexpected relationships were not included in the protocol. There was no identifiable influence of NS histology on ETP in either cohort (*p*=0.26). Antithrombin activity did not correlate with ETP in either cohort (*ρ*=‾0.06; *p*=0.47 and ‾0.23; *p*=0.28 for NEPTUNE and Columbus, respectively; **Figure S1**). In addition to ETP, several other parameters are derived from thrombin generation assays (i.e. lag-time, velocity index, peak thrombin, time to peak). In contrast to ETP, these other parameters were insignificantly or only weakly correlated with NS activity markers (data not shown).

**Table 2.** NEPTUNE Cohort Univariate Relationships with Endogenous Thrombin Potential.

|  | **B [95% CI]** | **p** |
| --- | --- | --- |
| Continuous Age (per year) | -15.59 [-27.45 to -3.72] | 0.0104 |
| Age (<18 vs. ≥18 years) | 887.19 [321.8 to 1452.59] | 0.0023 |
| Female vs. male | 199.46 [-347.04 to 745.96] | 0.4719 |
| Race: Black vs. non-Black | 469 [-99.62 to 1037.62] | 0.1052 |
| Hispanic vs. non-Hispanic | 428.28 [-284.5 to 1141.05] | 0.2369 |
| MN vs. FSGS | 597.91 [-120.91 to 1316.73] | 0.1023 |
| MCD vs. FSGS | 338.51 [-273.93 to 950.95] | 0.2764 |
| Duration of disease (months) | 1.4 [-2.55 to 5.34] | 0.4852 |
| Log-UP:C (g/g) | 265.39 [106.01 to 424.77] | 0.0013 |
| Plasma albumin (g/dl) | -656.73 [-957.24 to -356.21] | <.0001 |
| eGFR (ml/min) | 9.51 [1.88 to 17.13] | 0.0149 |
| Log-Total cholesterol | 2128.71 [1401.89 to 2855.53] | <.0001 |
| Log-HDL cholesterol | 1157.58 [413.53 to 1901.62] | 0.0025 |
| Log-LDL cholesterol | 1301.44 [763.5 to 1839.38] | <.0001 |
| Log-Triglycerides | 667.46 [196.28 to 1138.63] | 0.0058 |
| Treated with Lipid Modifying Agents | -32.31 [-592.69 to 528.07] | 0.9094 |
| Treated with Steroids | 853.24 [282.6 to 1423.87] | 0.0036 |
| Treated with any immunosuppression | 529.05 [-22.32 to 1080.43] | 0.0599 |
| Treated with RAAS-blockade | -572.74 [-1102.95 to -42.54] | 0.0344 |
| Alcohol use (ordinal) | -166.15 [-296.65 to -35.65] | 0.0130 |
| Tobacco use (ordinal) | -292 [-724.20 to 139.89] | 0.18 |
| Log-Antithrombin activity | -252.48 [-680.47 to 175.51] | 0.25 |

MN: membranous nephropathy, MCD: minimal change disease, FSGS: focal segmental glomerulosclerosis. UP:C: urine protein-to-creatinine ratio, eGFR: estimated glomerular filtration rate. HDL: high-density lipoprotein, LDL: low-density lipoprotein. RAAS: renin-angiotensin-aldosterone system.

**Fig. 2.**
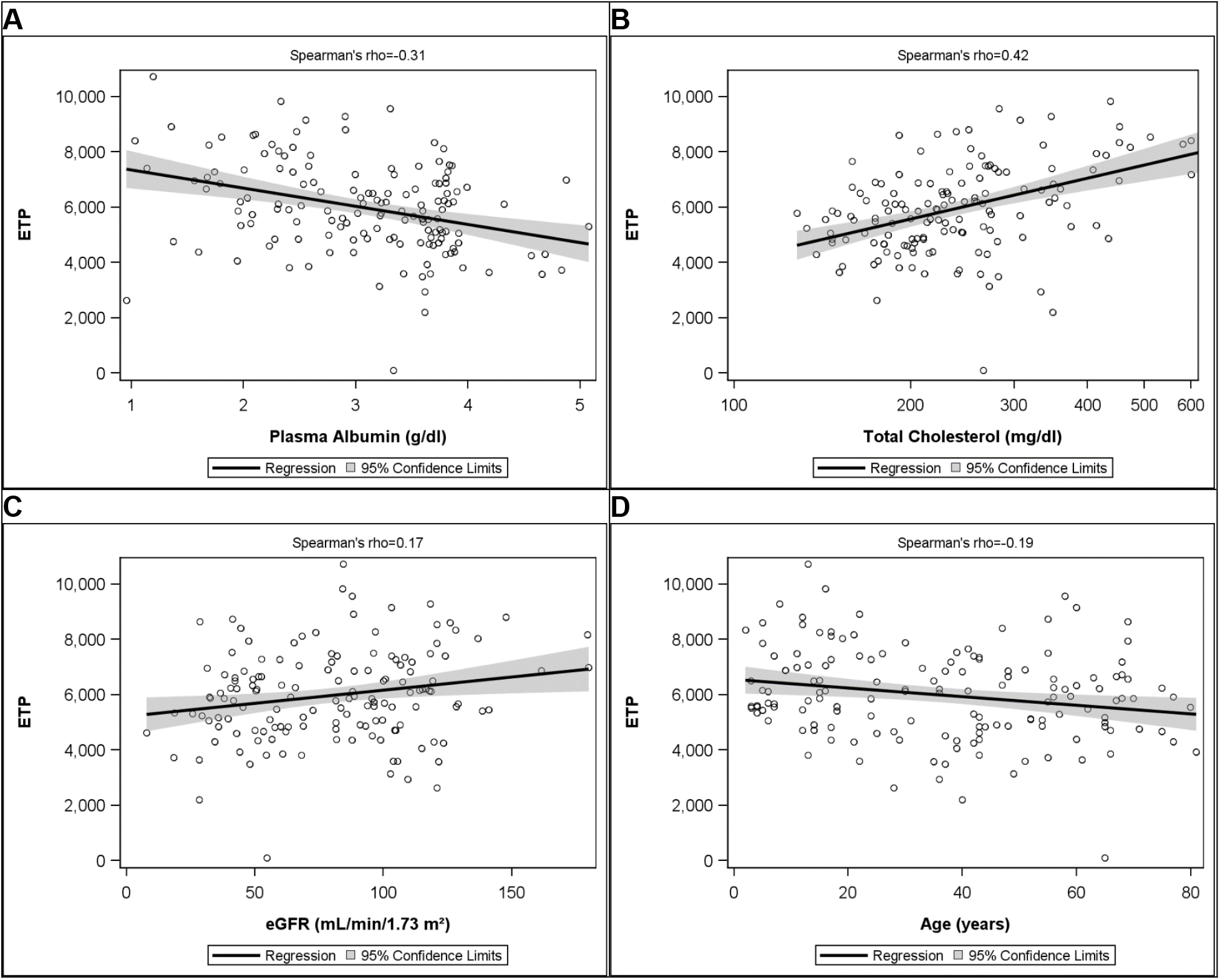
Univariate Relationships with Endogenous Thrombin Potential. In patients with nephrotic syndrome, endogenous thrombin potential (ETP; nM*min) is correlated with (**A**) plasma albumin, (**B**) total cholesterol, (**C**) estimated glomerular filtration rate (eGFR), and (**D**) Age.

**Fig. 3.**
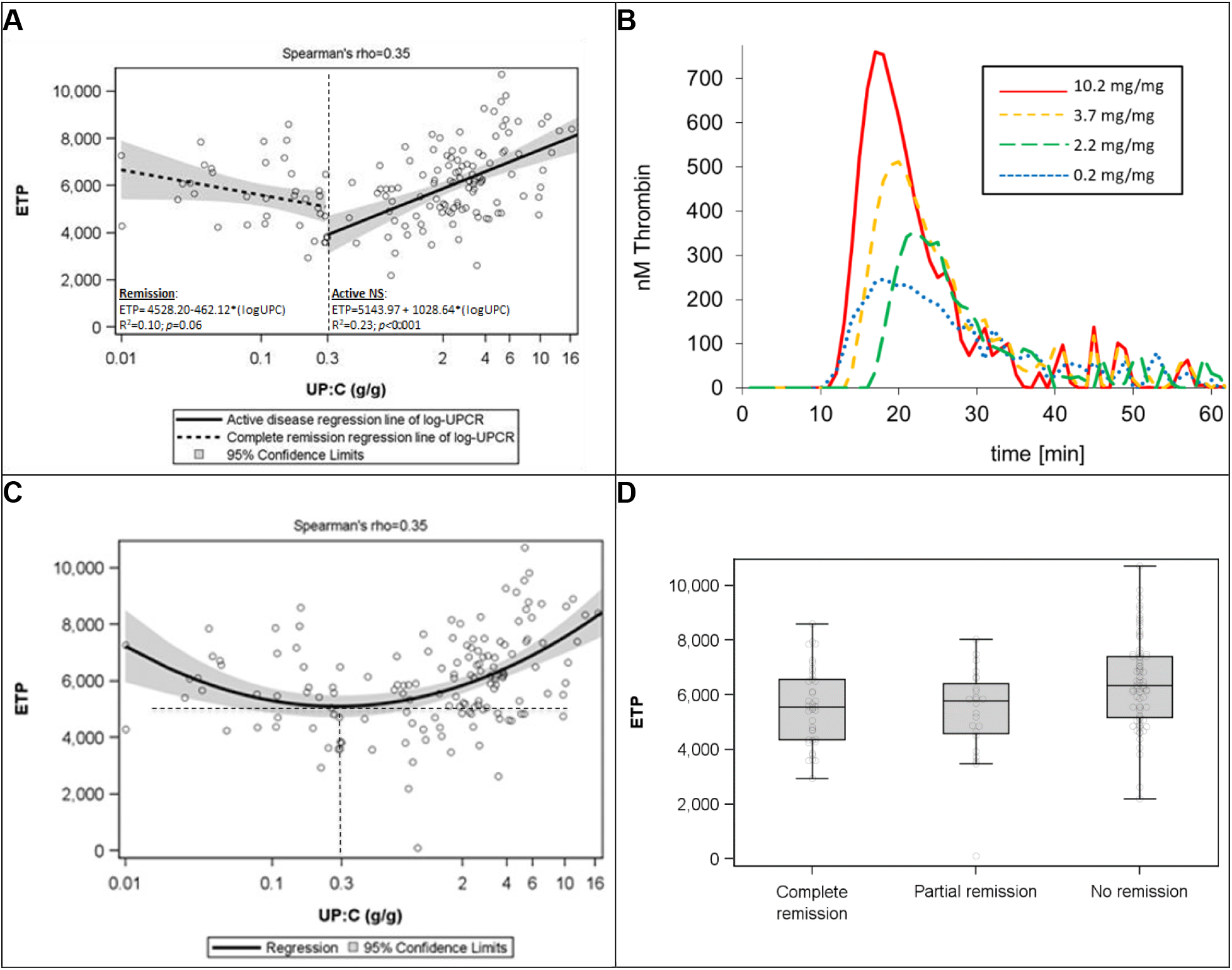
Relationship between Endogenous Thrombin Potential and Proteinuria is Dependent on Remission Status. (**A**) Endogenous thrombin potential (ETP; nM*min) is not significantly correlated with complete remission urinary protein-to creatinine ratio (UP:C) values (≤0.3 g/g creatinine) but significantly and positively correlated with UP:C in patients with active disease (>0.3 g/g creatinine). (**B**) Representative thrombin generation curves from four individual NEPTUNE patients representing the 1^st^, 2^nd^, 3^rd^, and 4^th^ UP:C quartiles of the NEPTUNE cohort. (**C**) Quadratic regression was utilized to mathematically analyze complex relationship between ETP and UP:C, demonstrating that the nadir ETP in the NEPTUNE cohort is nearly identical (0.28 g/g creatinine) to the complete remission UP:C cut-off (0.3 g/g creatinine). (**D**) ETP is significantly higher in NEPTUNE patients with active disease (no remission) than in those who have achieved complete or partial remission (*p* <0.001).

### Relationship between Endogenous Thrombin Potential and Proteinuria is Dependent on Remission Status

The UP:C scatterplot suggested a non-linear relationship between ETP and Log-UP:C (**Figure 3A**). Thus, the data were further analyzed after division into two groups: complete remission (CR; defined *a priori* for NEPTUNE as ≤0.3 g/g creatinine) and active disease (>0.3 g/g creatinine).^38^ This analysis revealed that the correlation was indirect and non-significant for patients in CR (*R*^2^=0.1; *p*=0.06), whereas there was a significant direct correlation among those with active disease (*R*^2^=0.23; *p*<0.001), suggesting an important interaction between ETP and remission status. When a quadratic regression formula was applied to the raw data the ETP nadir corresponded to UP:C 0.28 g/g creatinine (**Figure 3C**), mathematically confirming an interaction at a UP:C value that closely approximates the pre-defined CR threshold. Finally, when the subjects were grouped by remission status (CR vs. partial remission (PR) vs. no remission), those not in complete or partial remission had significantly higher ETP than either the CR or PR group (*p*<0.001; **Figure 3D**). Importantly, the observation that ETP was significantly and positively correlated with UP:C during active disease was also confirmed in the Columbus Cohort of newly-diagnosed NS patients (*ρ*=0.51; *p*=0.01).

### Disease Activity Biomarkers are Independently Predictive of Endogenous Thrombin Potential

Backward selection multivariable linear regression was used to construct an overall model of the relationship between ETP and NS disease activity (**Table 3**). All three NS activity biomarkers (UP:C, plasma albumin, and total cholesterol) were independently predictive of ETP, along with dichotomous age (children vs. adults). However, the other variables identified by univariate analysis were no longer predictive when incorporated into the multivariable model. The final model was significantly predictive of ETP (*R*^2^=0.38). The effect size of each variable included in the final model was estimated with the eta-squared (*η*^2^) function, wherein the sum of all *η*^2^ is equal to the overall correlation coefficient (*R*^2^).^39^ This analysis revealed that proteinuria had the largest influence on ETP (*η*^2^_Log-UP:C_+*η*^2^_Log-UP:C_^2^=*η*^2^ _proteinuria_=0.20), followed by total cholesterol (*η*^2^_total cholesterol_ =0.09), age (*η*^2^_age_ =0.07), and plasma albumin (*η*^2^_plasma albumin_=0.02). Separate models revealed that overall NS activity was significantly predictive of ETP in both adults (*R*^2^=0.27) and children (*R*^2^=0.57). When the models were reconstructed using the UP:C-defined remission interaction function rather than quadratic UP:C, they were similarly predictive with only minor changes in the correlation coefficients (Overall model: *R*^2^=0.40, Adults only: *R*^2^=0.29, Children only: *R*^2^=0.57; **Table S3**). In all models, UP:C had the greatest effect on ETP, followed by total cholesterol, with plasma albumin having the least influence.

**Table 3.** Disease Activity Biomarkers are Independently Predictive of Endogenous Thrombin Potential.

|  | **B [95% CI]** | **p** | **$\eta^2$** |
| --- | --- | --- | --- |
| **Overall ETP Model ($R^2=0.38$)** |  |  |  |
| Intercept | -1432.64 | --- | --- |
| Log-UP:C (g/g) | 332.28 [149.74 to 514.82] | 0.0004 | 0.07 |
| Log-UP:C<sup>2</sup> ([g/g]<sup>2</sup>) | 140.38 [65.26 to 215.51] | 0.0003 | 0.13 |
| Log-Total Cholesterol (mg/dl) | 1404 [687.44 to 2120.57] | 0.0002 | 0.09 |
| Plasma Albumin (g/dl) | -325.37 [-616.3 to -34.45] | 0.0287 | 0.02 |
| Age (<18 vs. ≥18 years) | 950.4 [461.01 to 1439.79] | 0.0002 | 0.07 |
| **Adult ETP Model ($R^2=0.27$)** |  |  |  |
| Intercept | -659.43 | --- | --- |
| Log-UP:C (g/g) | 363.8 [120.55 to 607.05] | 0.0038 | 0.07 |
| Log-UP:C<sup>2</sup> ([g/g]<sup>2</sup>) | 149 [52.04 to 245.96] | 0.0030 | 0.13 |
| Log-Total Cholesterol (mg/dl) | 1204.26 [282.11 to 2126.41] | 0.0110 | 0.06 |
| Plasma Albumin (g/dl) | -230.53 [-597.24 to 136.18] | 0.2152 | 0.01 |
| **Pediatric ETP Model ($R^2=0.57$)** |  |  |  |
| Intercept | -927.48 | --- | --- |
| Log-UP:C (g/g) | 291.48 [6.94 to 576.03] | 0.0449 | 0.20 |
| Log-UP:C<sup>2</sup> ([g/g]<sup>2</sup>) | 143.08 [19.36 to 266.79] | 0.0246 | 0.12 |
| Log-Total Cholesterol (mg/dl) | 1621.4 [371.66 to 2871.14] | 0.0124 | 0.19 |
| Plasma Albumin (g/dl) | -573.98 [-1113.73 to -34.24] | 0.0378 | 0.06 |

Multivariable regression using the quadratic urine protein-to-creatinine ratio (UP:C) formula reveals that proteinuria, total cholesterol, plasma albumin, and age are independently predictive of endogenous thrombin potential (ETP). The effect size of each variable toward the overall correlation of the model ( $R^2$ ) was estimated using the $\eta^2$ function. The total effect of proteinuria ( $\eta^2_{\text{Log-UP:C}} + \eta^2_{\text{Log-UP:C}^2} = \eta^2_{\text{proteinuria}}$ ) was 0.20 in both the overall and adult model whereas it was 0.32 in the pediatric model.

## Discussion

In this study of comprehensively phenotyped patients we found that hypercoagulopathy, as determined by ETP, is proportional to NS disease activity and confirmed this finding in a separate cohort.^40^ Importantly, NS-hypercoagulopathy improved commensurately with complete or partial remission. We also found that the acquired hyperlipidemia of NS has a previously unreported influence on NS-hypercoagulopathy. These data suggest that both proteinuria and hyperlipidemia exert greater influence on NS-hypercoagulopathy than albumin, which is presently the only clinically utilized biomarker.^15, 16^ In contrast to studies linking VTE-risk to both acquired antithrombin (AT) deficiency and NS histologic classification, we found no association between these variables and NS-hypercoagulopathy.^4-7^ Our multivariable models demonstrated that NS disease activity biomarkers were independently predictive of ETP and that the composite models were more predictive than the individual variables. These findings are expected to inform the design of studies needed to move toward evidence-based anticoagulant prophylaxis recommendations for patients with NS.

We have previously demonstrated that ETP is proportional to disease activity in NS animal models.^23^ The present data show that these pre-clinical observations are translatable to human NS, suggesting that these animal models may be utilized for robust pre-clinical investigations of NS-hypercoagulopathy. Recently, viscoelasticity assays have been used to characterize human NS-hypercoagulopathy.^21, 41^ Viscoelasticity was proportional to hypoalbuminemia, but not proteinuria, in MN patients whereas MCD patients were similar to healthy controls.^21^ However, in a subsequent MN study, the same laboratory reported viscoelastic changes proportional to proteinuria, serum albumin, triglycerides, platelet count, fibrinogen, and AT.^41^ While whole blood viscoelasticity assays thus offer a promising method for clinical NS-hypercoagulopathy research, they are difficult to apply to multicenter study settings because they require fresh whole blood samples, making interlaboratory standardization problematic.^42, 43^ Others have shown that ETP is elevated in human NS but the studies were underpowered to demonstrate a significant relationship with disease activity.^22^ Here we found that ETP was significantly related to NS activity and is thus a robust NS-hypercoagulopathy marker amenable to centralized laboratory testing using frozen and shipped samples. While this may simplify the analysis of hypercoagulopathy in multicenter NS studies, thrombin generation may not detect important contributions from cellular elements (e.g. platelets) that may be captured using whole blood viscoelasticity methods. Nonetheless, our animal model data suggest that the thrombin generation and viscoelastic methods may be interchangeable.^23^

Epidemiologic studies have shown that VTE-risk during NS is dependent upon both serum albumin and proteinuria, with albumin being the most widely studied.^10, 11, 17-20^ Thus, current guidelines suggest the use of serum albumin as a clinically relevant VTE-risk biomarker.^15^ As expected, ETP was dependent upon both proteinuria and albumin levels in our univariate analyses. Interestingly, total cholesterol was also independently predictive of ETP. Although hyperlipidemia is a well-described diagnostic feature of NS, it is generally not followed as a biomarker of NS disease activity.^3, 38^ However, hyperlipidemia may contribute to atherothrombotic complications of NS and has been associated with VTE-risk in post-menopausal women.^1, 44, 45^ Surprisingly, in this study, total cholesterol was more strongly associated with ETP status than was albumin. In MN patients, triglycerides (but not cholesterol) are associated with viscoelastic hypercoagulopathy which was ameliorated in patients taking statins.^41^ Similarly, statin use has been shown to both reduce VTE-risk in patients with NS and to reduce ETP in VTE patients without NS.^46, 47^ Moreover, cholesterol is known to enhance thrombin generation, an effect that is correctable with statin therapy.^48^ Very low-density lipoprotein (VLDL) has been shown to have the greatest effect on thrombin generation, followed by chylomicrons, and low-density lipoprotein (LDL).^49, 50^ However, oxidized LDL has a similar effect on thrombin generation as VLDL.^49^ Because all lipid components may be elevated in NS and were associated with ETP in this study, further study will be required to reveal which lipid components contribute most to thrombin generation during NS.^1, 44^

It has long been suggested that NS-hypercoagulopathy is due to urinary losses of AT resulting in intravascular AT depletion.^4-7^ Thus, it was not unexpected that 28% of the NEPTUNE samples had an AT activity <70%, a level which may be considered clinically relevant.^51^ We thus postulated that AT activity would be strongly correlated with ETP. To our surprise, however, we found no relationship between AT and ETP. This suggests that other, yet unknown, mechanisms drive ETP elevation during NS. As reviewed by our group and others, several other mechanisms (including hyperlipidemia) have been proposed,^4, 6^ but limited biorepository sample volume availability precluded further mechanistic exploration within the context of this study.

While NS, regardless of underlying histopathology, is associated with increased VTE risk, epidemiologic studies suggest that VTE is most prevalent amongst those with MN.^4, 13-15, 18, 19, 52^ It was thus surprising that ETP did not vary significantly by histopathology and thus does not explain the histopathologic variability in VTE predisposition. With the recent discovery that autoimmunity against phospholipase A2 receptor (PLA2R) or thrombospondin type-1 domain-containing 7A (THSD7A) are major mechanisms underlying MN, it is possible that disrupted PLA2R or THSD7A signaling are driving a prothrombotic vascular response.^53-57^ Local vascular responses might also explain the peculiar propensity for renal vein thrombosis during MN.^4^

Previous non-NS studies have demonstrated that ETP increases with age.^58, 59^ In contrast, we observed higher ETP in children than adults and no correlation with age within either age group. This difference may simply reflect sample size limitations, since we did not set out to study age as an EPT variable, or may be a function of NS pathology such that NS-hypercoagulopathy has a greater influence on ETP than does age. These data are limited by the lack of one or more confirmatory hypercoagulability assays, which were not possible given the biorepository sample volume restraints. Nonetheless, these data are congruent with previous single institution studies using viscoelastic methods.^21^ They are also congruent with the viscoelastic and ETP patterns we previously described in NS animal models.^23^ Moreover, we found similar patterns in the Columbus cohort which provides some evidence of internal validity. While this study was not designed or powered to determine ETP or NS activity biomarker thresholds predictive of thrombosis, appropriate power for such a study can now be extrapolated from these data. Future studies should thus be directed toward discerning VTE-risk thresholds that may guide the development of more effective and safer use of anticoagulant prophylaxis for patients with NS.

In summary, NS is a profoundly prothrombotic medical condition due, at least in part, to its acquired hypercoagulopathy. NS-hypercoagulopathy has now been shown to be proportional to disease activity using both viscoelastic and thrombin generation assays. The latter technique offers the opportunity to comprehensively study these associations within multicenter NS cohorts using a centralized laboratory to standardize analyses.^24^ It is hoped that these studies will lead to the development of evidence-based biomarker thresholds to guide appropriate, safe, and effective use of anticoagulant prophylaxis for patients with NS.

## Methods

### Study Subjects

#### NEPTUNE Cohort

The composition of the <u>Nep</u>hrotic Syndrome S<u>tu</u>dy <u>Ne</u>twork (NEPTUNE) has been described previously.^38, 40, 60^ The study is registered at clinicaltrials.gov (NCT01209000) and each subject (or their parent/guardian) provided written, informed consent prior to participation. Participants were enrolled from 24 clinical sites in North America with local IRB approval at each institution. Briefly, adults and children with proteinuria (≥500 mg/d on a 24 h urine sample or UP:C ≥0.5 g/g on spot urine) were eligible for enrolment at the time of a first clinically indicated renal biopsy. Histologic classification of disease subtype (MCD, FSGS, or MN) was confirmed by core pathology review.^61-63^ Because renal biopsy at presentation is not the standard of care in pediatrics, children (≤18 y) were also eligible for enrolment based upon the above proteinuria criteria alone. Exclusion criteria included life expectancy <6 months, prior solid organ transplantation, and kidney manifestations of systemic disease. For the purposes of the present study, participants with a recorded prescription for anticoagulant or antiplatelet medications were excluded, as were participants with a pre-enrolment history of VTE. Sodium citrate anticoagulated blood was collected from NEPTUNE participants using standard of care phlebotomy techniques at the time of enrolment and at follow-up intervals, processed to plasma within one hour, aliquoted, frozen at ‾80°C, and transferred to the Michigan Biobank. Corresponding phenotypic data for each patient’s sample was collected from the NEPTUNE Data Coordinating Center database (tranSMART). The pre-defined criteria for complete and partial remission utilized for NEPTUNE (UP:C ≤0.3 and UP:C <3.5 plus a ≥50% UP:C reduction, respectively) were applied where indicated.

#### Columbus Cohort

Because thrombin generation is easily confounded by pre-analytic variables that are not easily controlled using biorepository samples and because thrombin generation samples should ideally be collected into sodium citrate supplemented with corn trypsin inhibitor, a locally derived comparison cohort was recruited.^37, 64^ These patients were subject to the same eligibility criteria as the NEPTUNE participants, including exclusion for pre-enrolment VTE history, anticoagulant, or antiplatelet medications. This portion of the study was approved by the Nationwide Children’s Hospital Institutional Review Board (IRB12-00290) in reciprocity with The Ohio State University Wexner Medical Center IRB (neither institution is a NEPTUNE site). Written informed consent was obtained from each participant or parent/guardian. Blood was collected only at the time of enrolment, processed to plasma within one hour, and locally maintained at ‾80°C until analyzed. A simultaneous random urine sample was collected without preservatives, aliquoted, and frozen at ‾80°C for UP:C analysis.

### Phenotypic Data

UP:C assays were performed in a NEPTUNE central laboratory, and these values were extracted along with other phenotypic data. For the Columbus cohort, NS subtype was extracted from institutional pathology reports and UP:C was determined using spectrophotometric methods by Nationwide Children’s clinical laboratory.

### Plasma Samples

Aliquots (200 μL) of plasma isolated from blood collected into 0.105 M buffered sodium citrate tubes (0.32% final concentration citrate) were obtained from the NEPTUNE biorepository. For the Columbus cohort, blood was collected into final concentration 0.32% sodium citrate / 1.45 μM corn trypsin inhibitor (Haematologic Technologies Inc., Essex Junction, VT, USA) and platelet poor plasma (PPP) was prepared as previously described, aliquoted, and frozen at ‾80°C until further analysis.^23, 37^

### Thrombin Generation Assays

Thrombin generation assays (TGA) were performed using the Technothrombin TGA kit (Technoclone, Vienna, Austria) and TGA RC Low reagent using a Spectramax M2 fluorescent plate reader (Molecular Devices, Sunnyvale, California). The hemostatic activators in RC Low reagent consist of ≤5 pM recombinant tissue factor lipidated in ≤3.2 μM phospholipid micelles (final concentration <0.6 mM phospholipid).^65, 66^ Relative fluorescence units (RFU) were converted to thrombin generation curves using Technoclone Evaluation Software which also calculates endogenous thrombin potential (ETP). A previous study demonstrated that meaningful thrombin generation data could be derived from human plasma at dilutions up to 1:4 and TGA parameters do not change significantly with dilutions up to 1:1.^35, 67^ Thus, due to limited biorepository specimen volumes, TGA dilution studies were performed on NEPTUNE samples randomly selected to represent the full spectrum of proteinuria to determine the minimal necessary sample volume (**Figure S2**). These experiments demonstrated no differences in TGA parameters over the full range of dilutions studied (neat, 3:1, 2:1, and 1:1 plasma-to-buffer ratios). Therefore, to conserve available aliquot volumes, the 2:1 dilution was used for all analyses reported in this study. All TGA were performed in duplicate and reported as the mean ETP.

### Albumin and Antithrombin Assays

Plasma albumin was quantified using a bromocresol purple commercial assay kit according to the manufacturer’s instructions (BioAssay Systems, Hayward, CA).^68^ It should be noted that plasma albumin differs from serum albumin by sodium citrate dilution in a variable manner that is dependent on the patient’s hematocrit (not recorded) at the time of collection (9:1 whole blood to anticoagulant ratio). Antithrombin activity was determined as previously described, modified to use human pooled normal plasma for the standard curve.^23, 69^

### Statistical Analyses

Descriptive analyses were conducted on all participants using frequencies and percentages for categorical variables and medians and interquartile ranges for continuous variables. Descriptive statistics were also stratified by cohort within NEPTUNE. Correlations between ETP and NS variables were displayed using scatter plots and quantified with Spearman’s correlation coefficients (*ρ*). ETP was modeled using linear regression. The following covariates were tested as unadjusted predictors of ETP: age, sex, race, ethnicity, disease cohort, duration of disease, log-UP:C (transformed due to right-skewedness), plasma albumin, eGFR, antithrombin activity, total cholesterol, HDL, LDL, triglycerides, treatment with lipid modifying agents, steroids, any immunosuppressive, or RAAS blockade (each treatment category as separate categories coded as “treated vs. untreated”), alcohol use, and tobacco use. Any variable that was significant with unadjusted *p*<0.05 on univariate analysis entered a multivariable backwards selection process. There was little missing data and analyses were performed on a complete case basis. There was complete data capture for ETP, UP:C, plasma albumin, and eGFR. Three participants were missing centrally measured cholesterol values. Variables were removed from the model in descending order of *p*-value until all remaining variables were significant with *p*<0.05. All analyses were performed using SAS v9.4 (Cary, NC).

## Data Availability

Data are available via tranSMART data base according to the NEPTUNE ancillary study policy (see: https://www.rarediseasesnetwork.org/cms/neptune/Healthcare-Professionals/Ancillary).

https://neptune-study.org/

## Acknowledgments

We are indebted to the patient volunteers for their participation in the NEPTUNE and Columbus cohorts as well as the investigators and staff at each of the enrollment centers who made this study possible.

## Author contributions

SVP, BHR, WES, MK, and BAK enrolled patients and collected samples. APW and KJW performed the laboratory assays. APW, JPT, and BAK performed the data analyses and statistics. MTN, WES, MK, and BAK conceived the study. MK and BAK provided financial support. APW, JPT, and BAK wrote the paper. All authors refined the final draft.

## Addendum

NEPTUNE Enrolling Centers:

Cleveland Clinic, Cleveland, OH: J Sedor*, K Dell*, M Schachere#, J Negrey#

Children’s Hospital, Los Angeles, CA: K Lemley*, E Lim#

Children’s Mercy Hospital, Kansas City, MO: T Srivastava*, A Garrett#

Cohen Children’s Hospital, New Hyde Park, NY: C Sethna*, K Laurent #

Columbia University, New York, NY: G Appel*, M Toledo#

Duke University, Durham, NC: L Barisoni*

Emory University, Atlanta, GA: L Greenbaum*, C Wang**, C Kang#

Harbor-University of California Los Angeles Medical Center: S Adler*, C Nast*‡, J LaPage#

John H. Stroger Jr. Hospital of Cook County, Chicago, IL: A Athavale*, M Itteera

Johns Hopkins Medicine, Baltimore, MD: A Neu*, S Boynton#

Mayo Clinic, Rochester, MN: F Fervenza*, M Hogan**, J Lieske*, V Chernitskiy#

Montefiore Medical Center, Bronx, NY: F Kaskel*, N Kumar*, P Flynn#

NIDDK Intramural, Bethesda MD: J Kopp*, J Blake#

New York University Medical Center, New York, NY: H Trachtman*, O Zhdanova**, F Modersitzki#, S Vento#

Stanford University, Stanford, CA: R Lafayette*, K Mehta#

Temple University, Philadelphia, PA: C Gadegbeku*, D Johnstone**, S Quinn-Boyle#

University Health Network Toronto: D Cattran*, M Hladunewich**, H Reich**, P Ling#, M Romano#

University of Miami, Miami, FL: A Fornoni*, C Bidot#

University of Michigan, Ann Arbor, MI: M Kretzler*, D Gipson*, A Williams#, J LaVigne#

University of North Carolina, Chapel Hill, NC: V Derebail*, K Gibson*, A Froment#, S Grubbs#

University of Pennsylvania, Philadelphia, PA: L Holzman*, K Meyers**, K Kallem#, J Lalli#

University of Texas Southwestern, Dallas, TX: K Sambandam*, Z Wang#, M Rogers#

University of Washington, Seattle, WA: A Jefferson*, S Hingorani**, K Tuttle** §, M Bray #, M Kelton#, A Cooper# §

Wake Forest University Baptist Health, Winston-Salem, NC: B Freedman*, JJ Lin**

Data Analysis and Coordinating Center: M Kretzler, L Barisoni, C Gadegbeku, B Gillespie, D Gipson, L Holzman, L Mariani, M Sampson, J Troost, J Zee, E Herreshoff, S Li, C Lienczewski, J Liu, T Mainieri, M Wladkowski, A Williams

Digital Pathology Committee: Carmen Avila-Casado (UHN-Toronto), Serena Bagnasco (Johns Hopkins), Joseph Gaut (Washington U), Stephen Hewitt (National Cancer Institute), Jeff Hodgin (University of Michigan), Kevin Lemley (Children’s Hospital LA), Laura Mariani (University of Michigan), Matthew Palmer (U Pennsylvania), Avi Rosenberg (NIDDK), Virginie Royal (Montreal), David Thomas (University of Miami), Jarcy Zee (Arbor Research) Co-Chairs: Laura Barisoni (Duke University) and Cynthia Nast (Cedar Sinai)

*Principal Investigator; **Co-investigator; #Study Coordinator

‡Cedars-Sinai Medical Center, Los Angeles, CA

§Providence Medical Research Center, Spokane, WA

**Fig. S1.**
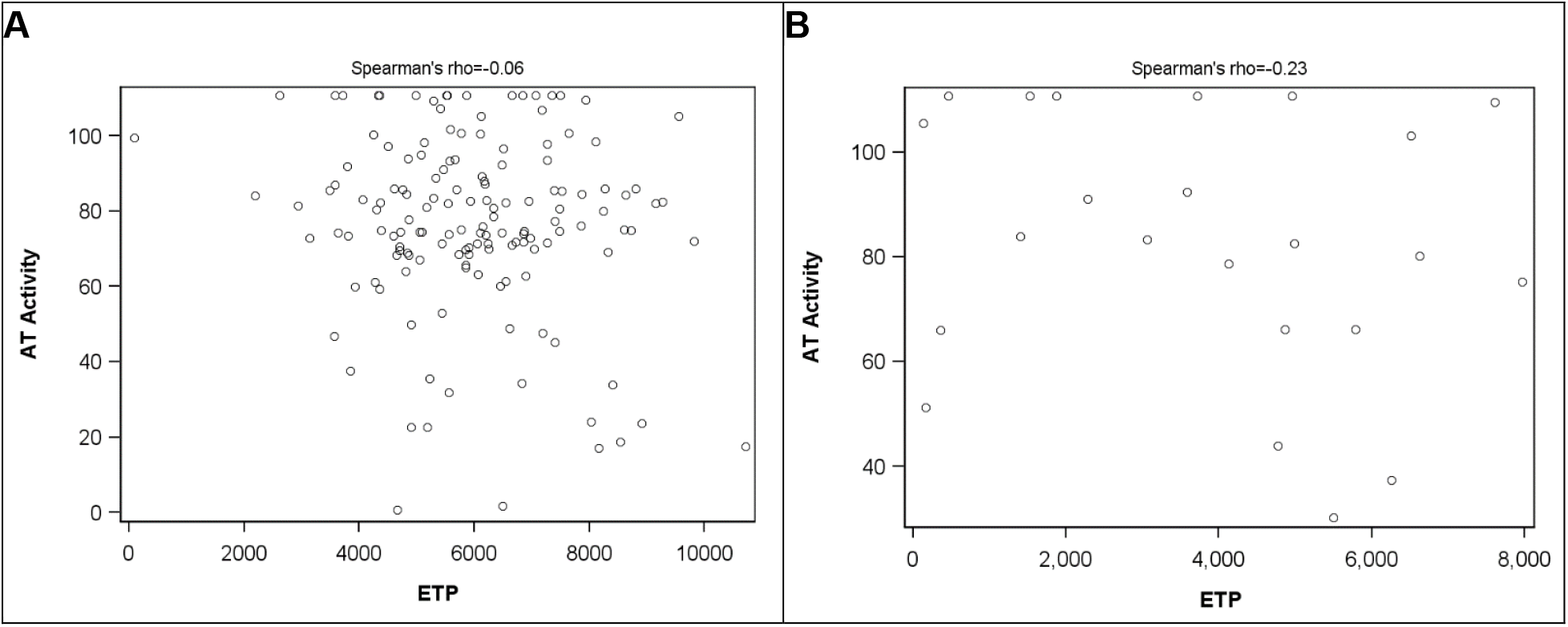
Antithrombin Activity does not Correlate with Endogenous Thrombin Potential. Univariate linear regression revealing no significant correlation between antithrombin (AT) activity (%) and endogenous thrombin potential (ETP; nM*min) in either the (**A**) NEPTUNE (*ρ*=‾0.06; *p*=0.47) or (**B**) Columbus (*ρ*=‾0.23; *p*=0.28) cohort.

**Fig. S2.**
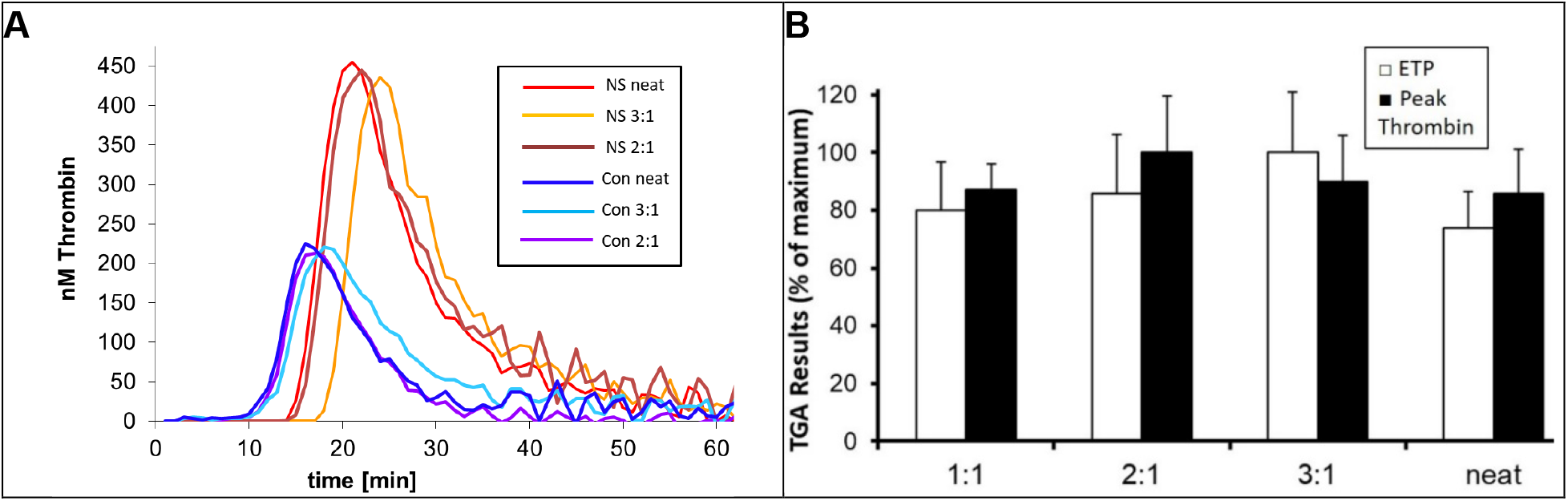
Thrombin Generation Assay Dilution Studies. (**A**) Representative thrombin generation assay (TGA) curves from one NS patient and one healthy volunteer plasma, analyzed with no dilution (neat), 3:1 (plasma:buffer ratio), and 2:1. (**B**) Mean ± SE values of endogenous thrombin potential (ETP) and peak thrombin from various plasma:buffer dilutions (*n*=6-10 plasma samples per dilution). Modest dilutions (3:1, 2:1, and 1:1) did not significantly alter either TGA parameter compared to neat. Due to the limited biorepository sample volumes available, the 2:1 dilution was used throughout the study.

**Table S1.**
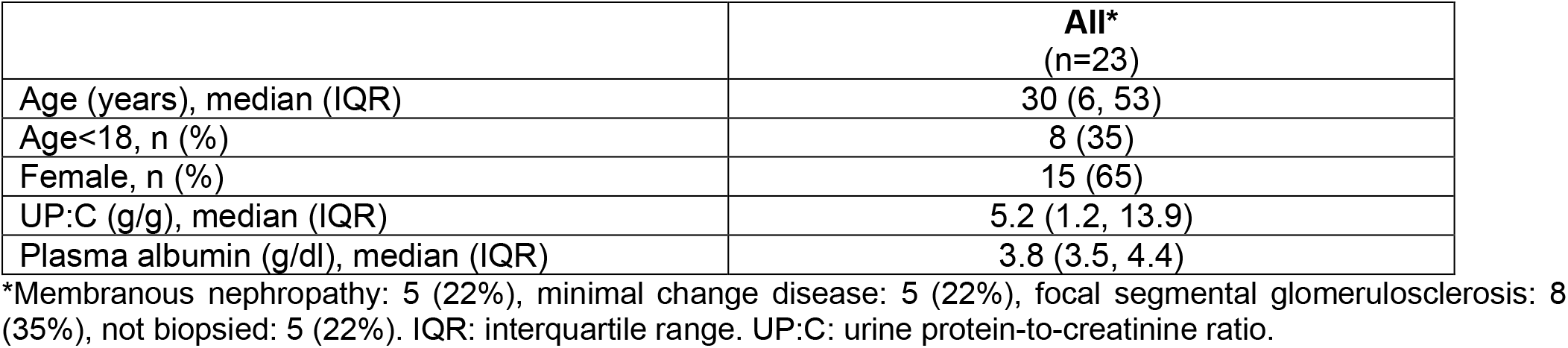
Demographics and Clinical Characteristics of the Columbus Cohort.

|  | **All\***<br>(n=23) |
| --- | --- |
| Age (years), median (IQR) | 30 (6, 53) |
| Age<18, n (%) | 8 (35) |
| Female, n (%) | 15 (65) |
| UP:C (g/g), median (IQR) | 5.2 (1.2, 13.9) |
| Plasma albumin (g/dl), median (IQR) | 3.8 (3.5, 4.4) |

\*Membranous nephropathy: 5 (22%), minimal change disease: 5 (22%), focal segmental glomerulosclerosis: 8 (35%), not biopsied: 5 (22%). IQR: interquartile range. UP:C: urine protein-to-creatinine ratio.

**Table S2.** Columbus Cohort Univariate Relationships with Endogenous Thrombin Potential.

|  | **B [95% CI]** | **p** |
| --- | --- | --- |
| Age (per year) | -25.57 [-72.6 to 21.46] | 0.2709 |
| Female vs. male | 348.09 [-1931.69 to 2627.88] | 0.7540 |
| FSGS vs. non-biopsied | -781.76 [-4435.46 to 2871.94] | 0.6593 |
| MCD vs. non-biopsied | -347.74 [-4223.08 to 3527.59] | 0.8530 |
| MN vs. non-biopsied | -837.88 [-4840.31 to 3164.55] | 0.6662 |
| Log-UP:C (g/g) | 853.1 [298.82 to 1407.39] | 0.0043 |
| Plasma albumin (g/dl) | -1760.93 [-3267.3 to -254.56] | 0.0241 |

FSGS: focal segmental glomerulosclerosis, MCD: minimal change disease, MN: membranous nephropathy. UP:C urine protein-to-creatinine ratio.

**Table S3.**
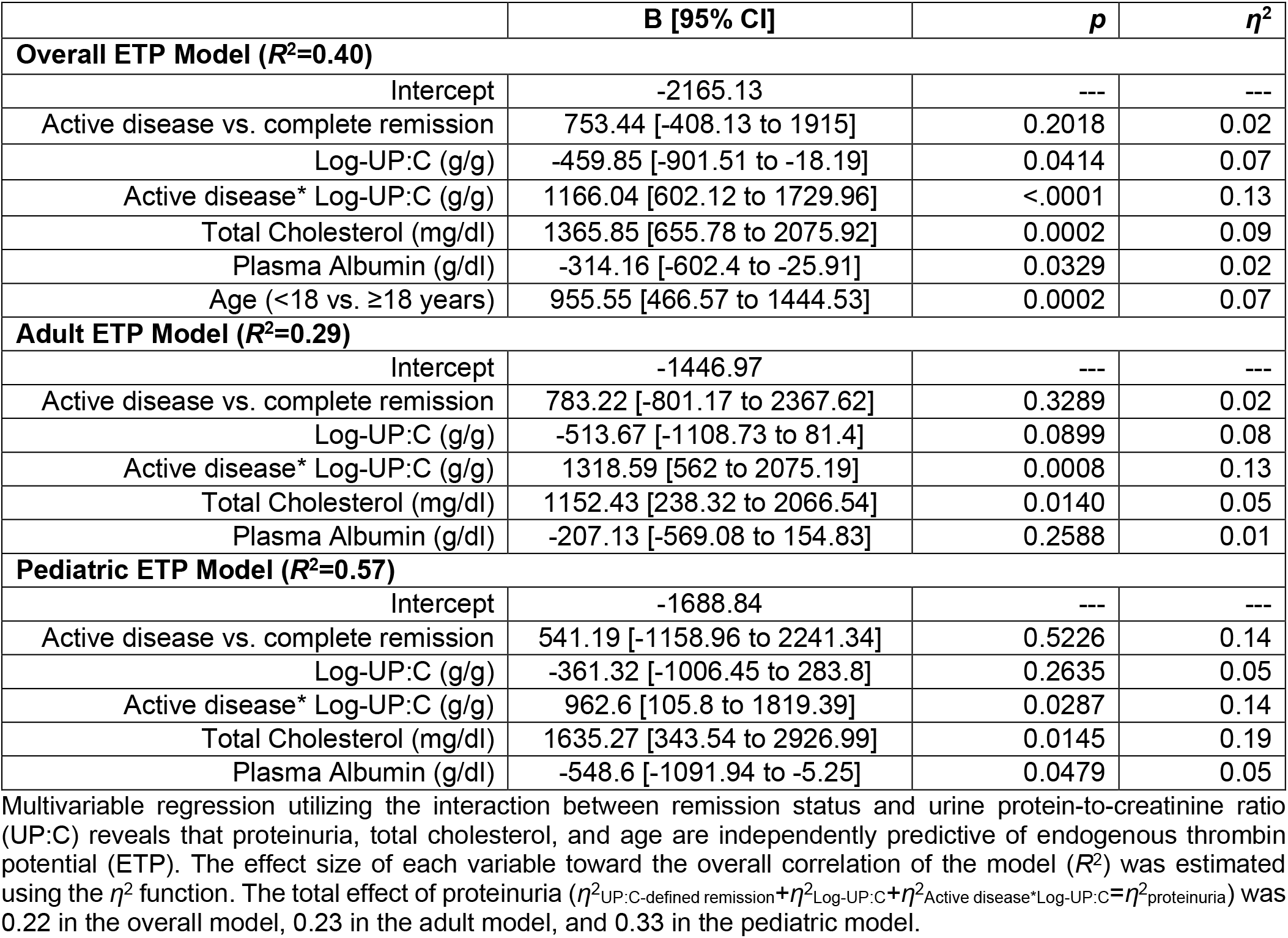
Disease Activity Biomarkers are Independently Predictive of Endogenous Thrombin Potential.

|  | **B [95% CI]** | **p** | **$\eta^2$** |
| --- | --- | --- | --- |
| **Overall ETP Model ($R^2=0.40$)** |  |  |  |
| Intercept | -2165.13 | --- | --- |
| Active disease vs. complete remission | 753.44 [-408.13 to 1915] | 0.2018 | 0.02 |
| Log-UP:C (g/g) | -459.85 [-901.51 to -18.19] | 0.0414 | 0.07 |
| Active disease* Log-UP:C (g/g) | 1166.04 [602.12 to 1729.96] | <.0001 | 0.13 |
| Total Cholesterol (mg/dl) | 1365.85 [655.78 to 2075.92] | 0.0002 | 0.09 |
| Plasma Albumin (g/dl) | -314.16 [-602.4 to -25.91] | 0.0329 | 0.02 |
| Age (<18 vs. ≥18 years) | 955.55 [466.57 to 1444.53] | 0.0002 | 0.07 |
| **Adult ETP Model ($R^2=0.29$)** |  |  |  |
| Intercept | -1446.97 | --- | --- |
| Active disease vs. complete remission | 783.22 [-801.17 to 2367.62] | 0.3289 | 0.02 |
| Log-UP:C (g/g) | -513.67 [-1108.73 to 81.4] | 0.0899 | 0.08 |
| Active disease* Log-UP:C (g/g) | 1318.59 [562 to 2075.19] | 0.0008 | 0.13 |
| Total Cholesterol (mg/dl) | 1152.43 [238.32 to 2066.54] | 0.0140 | 0.05 |
| Plasma Albumin (g/dl) | -207.13 [-569.08 to 154.83] | 0.2588 | 0.01 |
| **Pediatric ETP Model ($R^2=0.57$)** |  |  |  |
| Intercept | -1688.84 | --- | --- |
| Active disease vs. complete remission | 541.19 [-1158.96 to 2241.34] | 0.5226 | 0.14 |
| Log-UP:C (g/g) | -361.32 [-1006.45 to 283.8] | 0.2635 | 0.05 |
| Active disease* Log-UP:C (g/g) | 962.6 [105.8 to 1819.39] | 0.0287 | 0.14 |
| Total Cholesterol (mg/dl) | 1635.27 [343.54 to 2926.99] | 0.0145 | 0.19 |
| Plasma Albumin (g/dl) | -548.6 [-1091.94 to -5.25] | 0.0479 | 0.05 |

Multivariable regression utilizing the interaction between remission status and urine protein-to-creatinine ratio (UP:C) reveals that proteinuria, total cholesterol, and age are independently predictive of endogenous thrombin potential (ETP). The effect size of each variable toward the overall correlation of the model ( $R^2$ ) was estimated using the $\eta^2$ function. The total effect of proteinuria ( $\eta^2_{UP:C\text{-defined remission}} + \eta^2_{Log-UP:C} + \eta^2_{Active\ disease*Log-UP:C} = \eta^2_{proteinuria}$ ) was 0.22 in the overall model, 0.23 in the adult model, and 0.33 in the pediatric model.

## References

1. Agrawal S, Zaritsky JJ, Fornoni A, et al. Dyslipidaemia in nephrotic syndrome: mechanisms and treatment. Nat Rev Nephrol 2018; 14: 57–70.

2. Eddy AA, Symons JM. Nephrotic syndrome in childhood. Lancet 2003; 362: 629–639.

3. Orth SR, Ritz E. The nephrotic syndrome. The New England journal of medicine 1998; 338: 1202–1211.

4. Kerlin BA, Ayoob R, Smoyer WE. Epidemiology and pathophysiology of nephrotic syndrome-associated thromboembolic disease. Clinical journal of the American Society of Nephrology : CJASN 2012; 7: 513–520.

5. Loscalzo J. Venous thrombosis in the nephrotic syndrome. The New England journal of medicine 2013; 368: 956–958.

6. Schlegel N. Thromboembolic risks and complications in nephrotic children. Seminars in thrombosis and hemostasis 1997; 23: 271–280.

7. Singhal R, Brimble KS. Thromboembolic complications in the nephrotic syndrome: pathophysiology and clinical management. Thrombosis research 2006; 118: 397–407.

8. Lee T, Biddle AK, Lionaki S, et al. Personalized prophylactic anticoagulation decision analysis in patients with membranous nephropathy. Kidney international 2014; 85: 1412–1420.

9. Cushman M. Epidemiology and risk factors for venous thrombosis. Semin Hematol 2007; 44: 62–69.

10. Mahmoodi BK, ten Kate MK, Waanders F, et al. High absolute risks and predictors of venous and arterial thromboembolic events in patients with nephrotic syndrome: results from a large retrospective cohort study. Circulation 2008; 117: 224–230.

11. Kerlin BA, Blatt NB, Fuh B, et al. Epidemiology and risk factors for thromboembolic complications of childhood nephrotic syndrome: a Midwest Pediatric Nephrology Consortium (MWPNC) study. The Journal of pediatrics 2009; 155: 105-110, 110 e101.

12. Kerlin BA, Haworth K, Smoyer WE. Venous thromboembolism in pediatric nephrotic syndrome. Pediatric nephrology 2013.

13. Glassock RJ. Prophylactic anticoagulation in nephrotic syndrome: a clinical conundrum. J Am Soc Nephrol 2007; 18: 2221–2225.

14. Sarasin FP, Schifferli JA. Prophylactic oral anticoagulation in nephrotic patients with idiopathic membranous nephropathy. Kidney international 1994; 45: 578–585.

15. KDIGO Clinical Practice Guideline for Glomerulonephritis. Kidney Int Suppl 2012; 2: 139–274.

16. Derebail VK, Rheault MN, Kerlin BA. Role of Direct Oral Anticoagulants in Patients with Kidney Disease. Kidney international In Press Journal Pre-Proof, 2019.

17. Kumar S, Chapagain A, Nitsch D, et al. Proteinuria and hypoalbuminemia are risk factors for thromboembolic events in patients with idiopathic membranous nephropathy: an observational study. BMC nephrology 2012; 13: 107.

18. Lionaki S, Derebail VK, Hogan SL, et al. Venous thromboembolism in patients with membranous nephropathy. Clinical journal of the American Society of Nephrology : CJASN 2012; 7: 43–51.

19. Bellomo R, Atkins RC. Membranous nephropathy and thromboembolism: is prophylactic anticoagulation warranted? Nephron 1993; 63: 249–254.

20. Barbour SJ, Greenwald A, Djurdjev O, et al. Disease-specific risk of venous thromboembolic events is increased in idiopathic glomerulonephritis. Kidney international 2012; 81: 190–195.

21. Huang MJ, Wei RB, Wang ZC, et al. Mechanisms of hypercoagulability in nephrotic syndrome associated with membranous nephropathy as assessed by thromboelastography. Thrombosis research 2015; 136: 663–668.

22. Mahmoodi BK, Mulder AB, Waanders F, et al. The impact of antiproteinuric therapy on the prothrombotic state in patients with overt proteinuria. Journal of thrombosis and haemostasis : JTH 2011; 9: 2416–2423.

23. Kerlin BA, Waller AP, Sharma R, et al. Disease Severity Correlates with Thrombotic Capacity in Experimental Nephrotic Syndrome. J Am Soc Nephrol 2015; 26: 3009–3019.

24. Brummel-Ziedins KE, Wolberg AS. Global assays of hemostasis. Curr Opin Hematol 2014; 21: 395–403.

25. Lane DA, Philippou H, Huntington JA. Directing thrombin. Blood 2005; 106: 2605–2612.

26. Hemker HC, Al Dieri R, De Smedt E, et al. Thrombin generation, a function test of the haemostatic-thrombotic system. Thromb Haemost 2006; 96: 553–561.

27. Besser M, Baglin C, Luddington R, et al. High rate of unprovoked recurrent venous thrombosis is associated with high thrombin-generating potential in a prospective cohort study. Journal of thrombosis and haemostasis : JTH 2008; 6: 1720–1725.

28. Hron G, Kollars M, Binder BR, et al. Identification of patients at low risk for recurrent venous thromboembolism by measuring thrombin generation. Jama 2006; 296: 397–402.

29. van Hylckama Vlieg A, Baglin CA, Luddington R, et al. The risk of a first and a recurrent venous thrombosis associated with an elevated D-dimer level and an elevated thrombin potential: results of the THE-VTE study. Journal of thrombosis and haemostasis : JTH 2015; 13: 1642–1652.

30. Emani S, Zurakowski D, Baird CW, et al. Hypercoagulability panel testing predicts thrombosis in neonates undergoing cardiac surgery. American journal of hematology 2014; 89: 151–155.

31. Emani S, Zurakowski D, Baird CW, et al. Hypercoagulability markers predict thrombosis in single ventricle neonates undergoing cardiac surgery. The Annals of thoracic surgery 2013; 96: 651–656.

32. Sonnevi K, Tchaikovski SN, Holmstrom M, et al. Obesity and thrombin-generation profiles in women with venous thromboembolism. Blood coagulation & fibrinolysis : an international journal in haemostasis and thrombosis 2013; 24: 547–553.

33. Ay C, Dunkler D, Simanek R, et al. Prediction of venous thromboembolism in patients with cancer by measuring thrombin generation: results from the Vienna Cancer and Thrombosis Study. Journal of clinical oncology : official journal of the American Society of Clinical Oncology 2011; 29: 2099–2103.

34. Eichinger S, Hron G, Kollars M, et al. Prediction of recurrent venous thromboembolism by endogenous thrombin potential and D-dimer. Clinical chemistry 2008; 54: 2042–2048.

35. van Hylckama Vlieg A, Christiansen SC, Luddington R, et al. Elevated endogenous thrombin potential is associated with an increased risk of a first deep venous thrombosis but not with the risk of recurrence. British journal of haematology 2007; 138: 769–774.

36. Tripodi A, Martinelli I, Chantarangkul V, et al. The endogenous thrombin potential and the risk of venous thromboembolism. Thrombosis research 2007; 121: 353–359.

37. Dargaud Y, Luddington R, Gray E, et al. Standardisation of thrombin generation test--which reference plasma for TGT? An international multicentre study. Thrombosis research 2010; 125: 353–356.

38. Gipson DS, Troost JP, Lafayette RA, et al. Complete Remission in the Nephrotic Syndrome Study Network. Clinical journal of the American Society of Nephrology : CJASN 2016; 11: 81–89.

39. Cohen J. Eta-Squared and Partial Eta-Squared in Fixed Factor Anova Designs. Educ Psychol Meas 1973; 33: 107–112.

40. Gadegbeku CA, Gipson DS, Holzman LB, et al. Design of the Nephrotic Syndrome Study Network (NEPTUNE) to evaluate primary glomerular nephropathy by a multidisciplinary approach. Kidney international 2013; 83: 749–756.

41. Huang MJ, Wei RB, Li QP, et al. Hypercoagulable state evaluated by thromboelastography in patients with idiopathic membranous nephropathy. J Thromb Thrombolysis 2016; 41: 321–327.

42. Chitlur M, Lusher J. Standardization of thromboelastography: values and challenges. Seminars in thrombosis and hemostasis 2010; 36: 707–711.

43. Chitlur M, Sorensen B, Rivard GE, et al. Standardization of thromboelastography: a report from the TEG-ROTEM working group. Haemophilia 2011; 17: 532–537.

44. Vaziri ND. Disorders of lipid metabolism in nephrotic syndrome: mechanisms and consequences. Kidney international 2016; 90: 41–52.

45. Heit JA, Spencer FA, White RH. The epidemiology of venous thromboembolism. J Thromb Thrombolysis 2016; 41: 3–14.

46. Resh M, Mahmoodi BK, Navis GJ, et al. Statin use in patients with nephrotic syndrome is associated with a lower risk of venous thromboembolism. Thrombosis research 2011; 127: 395–399.

47. Orsi FA, Biedermann JS, Kruip M, et al. Rosuvastatin use reduces thrombin generation potential in patients with venous thromboembolism: a randomized controlled trial. Journal of thrombosis and haemostasis : JTH 2019; 17: 319–328.

48. Aoki I, Aoki N, Kawano K, et al. Platelet-dependent thrombin generation in patients with hyperlipidemia. J Am Coll Cardiol 1997; 30: 91–96.

49. Rota S, McWilliam NA, Baglin TP, et al. Atherogenic lipoproteins support assembly of the prothrombinase complex and thrombin generation: modulation by oxidation and vitamin E. Blood 1998; 91: 508–515.

50. Moyer MP, Tracy RP, Tracy PB, et al. Plasma lipoproteins support prothrombinase and other procoagulant enzymatic complexes. Arterioscler Thromb Vasc Biol 1998; 18: 458–465.

51. Patnaik MM, Moll S. Inherited antithrombin deficiency: a review. Haemophilia 2008; 14: 1229–1239.

52. Nickolas TL, Radhakrishnan J, Appel GB. Hyperlipidemia and thrombotic complications in patients with membranous nephropathy. Semin Nephrol 2003; 23: 406–411.

53. Tomas NM, Beck LH, Jr., Meyer-Schwesinger C, et al. Thrombospondin type-1 domain-containing 7A in idiopathic membranous nephropathy. The New England journal of medicine 2014; 371: 2277–2287.

54. Beck LH, Jr., Bonegio RG, Lambeau G, et al. M-type phospholipase A2 receptor as target antigen in idiopathic membranous nephropathy. The New England journal of medicine 2009; 361: 11–21.

55. Si NV, Fujioka D, Watanabe K, et al. Phospholipase A2 Receptor Gene Polymorphisms Alter its Functions and Present a Genetic Risk of an Increased Intima-Media Thickness of the Carotid Artery. J Atheroscler Thromb 2016; 23: 1227–1241.

56. Kuo MW, Wang CH, Wu HC, et al. Soluble THSD7A is an N-glycoprotein that promotes endothelial cell migration and tube formation in angiogenesis. PLoS One 2011; 6: e29000.

57. Wang CH, Su PT, D. Xy, et al. Thrombospondin type I domain containing 7A (THSD7A) mediates endothelial cell migration and tube formation. J Cell Physiol 2010; 222: 685–694.

58. Haidl H, Cimenti C, Leschnik B, et al. Age-dependency of thrombin generation measured by means of calibrated automated thrombography (CAT). Thromb Haemost 2006; 95: 772–775.

59. Wu J, Zhao HR, Zhang HY, et al. Thrombin generation increasing with age and decreasing with use of heparin indicated by calibrated automated thrombogram conducted in Chinese. Biomed Environ Sci 2014; 27: 378–384.

60. Sethna CB, Meyers KEC, Mariani LH, et al. Blood Pressure and Visit-to-Visit Blood Pressure Variability Among Individuals With Primary Proteinuric Glomerulopathies. Hypertension 2017; 70: 315–323.

61. Mariani LH, Martini S, Barisoni L, et al. Interstitial fibrosis scored on whole-slide digital imaging of kidney biopsies is a predictor of outcome in proteinuric glomerulopathies. Nephrology, dialysis, transplantation : official publication of the European Dialysis and Transplant Association - European Renal Association 2018; 33: 310–318.

62. Barisoni L, Troost JP, Nast C, et al. Reproducibility of the NEPTUNE descriptor-based scoring system on whole-slide images and histologic and ultrastructural digital images. Modern pathology : an official journal of the United States and Canadian Academy of Pathology, Inc 2016; 29: 671–684.

63. Barisoni L, Nast CC, Jennette JC, et al. Digital pathology evaluation in the multicenter Nephrotic Syndrome Study Network (NEPTUNE). Clinical journal of the American Society of Nephrology : CJASN 2013; 8: 1449–1459.

64. Favaloro EJ, Funk DM, Lippi G. Pre-analytical Variables in Coagulation Testing Associated With Diagnostic Errors in Hemostasis. Laboratory Medicine 2012; 43: 1–10.

65. Maji D, Nayak L, Martin J, et al. A novel, point-of-care, whole-blood assay utilizing dielectric spectroscopy is sensitive to coagulation factor replacement therapy in haemophilia A patients. Haemophilia 2019; 25: 885–892.

66. Konigsbrugge O, Koder S, Riedl J, et al. A new measure for in vivo thrombin activity in comparison with in vitro thrombin generation potential in patients with hyper- and hypocoagulability. Clin Exp Med 2017; 17: 251–256.

67. Chandler WL, Roshal M. Optimization of plasma fluorogenic thrombin-generation assays. Am J Clin Pathol 2009; 132: 169–179.

68. Ueno T, Hirayama S, Ito M, et al. Albumin concentration determined by the modified bromocresol purple method is superior to that by the bromocresol green method for assessing nutritional status in malnourished patients with inflammation. Ann Clin Biochem 2013; 50: 576–584.

69. Odegard OR, Lie M, Abildgaard U. Heparin cofactor activity measured with an amidolytic method. Thrombosis research 1975; 6: 287–294.

